# Optimising the mirror illusion during mirror therapy: evidence from unimpaired individuals

**DOI:** 10.1101/2024.11.16.24317419

**Authors:** J.M. Kim, F.O. Challis, C.Y. Koo, C.L. Leung, L.H. Lo, S.H. Yeo, T.D. Punt

## Abstract

Patients recovering from hemiparetic stroke have been shown to benefit from mirror therapy in terms of improving their motor function. The clinical improvement in motor function may differ depending on the mirror therapy protocol used. Previous studies have shown that four parameters are influential: the size of the mirror (large and small), manipulation of objects (with or without), the complexity of action (simple and complex), and movement execution (unilateral and bilateral). We examined the impact of these parameters on the subjective quality (believability) of the mirror illusion in unimpaired participants. Forty healthy participants completed 16 different combinations of the four parameters during mirror visual feedback. Participants rated each trial for its level of *believability* on a 10-point Likert scale. A repeated measures ANOVA was used to examine the data. The large mirror consistently elicited higher ratings than the small mirror. And while bimanual movements generally elicited higher ratings than unimanual movements, ratings for bimanual movements were significantly reduced when participants made complex movements with objects. We attributed these results to the congruency of multisensory information. Conditions that elicit congruency between illusory information and other sensory inputs appear to maximise believability over the illusory hand. The findings of this study reveal the parameters maximising illusion believability in unimpaired participants and have implications for optimal mirror therapy conditions in patients’ groups.

## Introduction

Mirror therapy has been found to be an effective intervention for the rehabilitation of paretic limbs following stroke (Altschuler et al., 1999). Most commonly employed as an intervention for the upper limb, evidence demonstrates that it improves motor impairment and function of the paretic limb (Thieme et al., 2018). While multiple clinical trials have been completed, the precise mechanism of mirror therapy remains uncertain. Furthermore, many different protocols exist, and it remains unclear what the optimal approach may be.

Morkisch et al. (2019) conducted a meta-analysis examining the impact of different mirror therapy protocols on the effectiveness of the intervention. Their analysis included all 32 studies that contributed to the earlier systematic review and had reported motor impairment and function data (Thieme et al., 2018). The impact of three components of the intervention was examined: (i) Mirror Size; small vs large mirror, (ii) Movement Execution; unimanual vs bimanual movement, and (iii) Movement Type; use of an object or not.

Data from their meta-analysis suggested mirror therapy was most effective when a large (rather than a small) mirror was used. Following the definition used by Kim and colleagues (2017), they classified the size of the mirror as a large mirror when it reaches eye level (50×40cm). The use of larger mirrors may help to make the illusion process more immersive (Ramachandran & Rogers-Ramachandran, 2019); more effectively obscuring the limb behind the mirror and perhaps eliciting enhanced attention to the hand reflected in the mirror. As the authors highlight, larger mirrors allow more of the limb to be reflected and may, therefore, facilitate greater adaptation (Morkisch et al., 2017).

Morkisch et al.’s (2019) meta-analysis also showed that encouraging patients to make only unimanual movements with the unaffected arm only (unimanual execution) was more effective than instructing patients to move both limbs together (bimanual execution). This finding is intriguing, not only because it differs from the initially proposed protocol for mirror therapy in individuals with hemiparetic stroke, but also because it discourages movement in the limb that the intervention is designed to improve function in. When Altschuler et al. (1999) first proposed mirror therapy for hemiparetic stroke, they suggested patients should move both arms together and symmetrically. Patients were encouraged to move the impaired limb hidden behind the mirror as much as possible. The results of the meta-analysis (Morkisch et al., 2019), therefore, seem counterintuitive as the recommendation removes any actual physical training of the impaired limb from the intervention. Rather than focusing on this, Morkisch et al. (2019) suggest that the weaker effect found for bimanual execution was due to the bilateral dispersion of attention during therapy.

Finally, Morkisch et al. (2019)’s meta-analysis demonstrated that effectiveness was reduced when the movements performed involved object manipulation; i.e. mirror therapy was more effective when movements were made in the absence of objects. Tasks that involve the manipulation of objects are often described as *task-oriented* (Arya et al., 2015; Higgins et al., 2006; Michaelsen et al., 2006); and task-oriented practice is typically recommended for upper limb rehabilitation in individuals following stroke (Bravi & Ellen Stoykov, 2007; da Silva et al., 2020; Veerbeek et al., 2014). Here again then we see that data from mirror therapy research is at odds with normal guidance; i.e. movements performed during mirror therapy should not involve objects in the normal task-oriented manner. Perhaps manipulating an object while receiving visual feedback in the mirror shifts attentional focus predominantly toward the ‘seen’ hand. This shift may occur because the visual feedback enhances the salience of the ‘seen’ hand, directing attention away from the illusory hand and reinforcing the sense of control over the movements of the ‘seen’ hand.

The potential negative impact of object manipulation was highlighted by Bai et al. (2019) who reported that movement-based mirror therapy improved motor impairment more than task-based mirror therapy in subacute stroke survivors. In this study, the movement-based group were instructed to make simple joint movements without an object, such as joint flexion and extension, gripping/releasing and finger tapping, while the task-based group were instructed to make relatively complex movements with objects, such as transferring cubes, placing pegs in holes and turning over paper cards. However, when comparing the tasks assigned to the two groups, the presence or absence of an object was not the only difference. Rather, the complexity of activities for the task-based group was far greater and this component may be of relevance too.

In all cases, the parameters highlighted by Morkisch et al. (2019) appear to have a significant impact on the quality of the illusory experience during mirror therapy; and perhaps optimising this experience is directly related to the effectiveness of the intervention. The concept of illusory information quality has been recognised as a cornerstone of mirror therapy since its inception (Ramachandran et al., 1995). McCabe (2011) emphasised that fully believing in the illusory limb could be critical to the success of the intervention. Additionally, Rowe et al. (2019) provided a more intuitive illustration of the illusory experience’s effects. Their study examined how task complexity affected the illusory experience, using a “task realism” scale across twenty-five different bimanual tasks. The findings revealed that participants rated simple movements without object manipulation as the most realistic.

As might be expected, illusion strength appears to depend on the congruency of sensory information. When sensory information from different modalities is congruent, sensory inputs are strengthened and facilitate behavioural responses (Alais et al., 2010). In contrast, where conflicts in sensory information occur, this appears to threaten the illusion (Wittkopf et al., 2019). Any incongruence with the illusory visual information may, therefore, undermine the illusion (Synofzik et al., 2006). A crossmodal illusion occurs when the senses received from one modality affect the senses received from other modalities, providing coherence to the ongoing perceptual experience (Bolognini et al., 2015). In the case of mirror therapy, visual information obtained through the mirror illusion affects the proprioceptive (Snijders et al., 2007) or tactile information (Katsuyama et al., 2018) from the unseen hand. As a result of multisensory integration, the presence of a mirror biases perception toward the (visual) illusory information (Holmes & Spence, 2005; Holmes et al., 2004).

The contribution of the crossmodal illusion during mirror visual feedback can be indicated through the investigation of the sense of embodiment (Ehrsson, 2020; Ernst & Bülthoff, 2004; Wittkopf et al., 2019). The sense of embodiment indicates how much one believes the hand reflected in the mirror as one’s own unseen hand. Sense of embodiment is typically considered to encompass three subcomponents. This includes whether the mirror image appears to be yours (ownership), whether you feel that the moving mirror image is under your control (agency), and whether you feel that the mirror image represents the location of the unseen hand (location) (Longo et al., 2008). However, embodiment can be simply investigated as a sense of realism (Rowe et al., 2019) or a peculiarity (Fink et al., 1999) in the mirror image during mirror visual feedback.

Beyond simply feeling that the image in the mirror is yours (ownership), the participant’s believability more intuitively judges the embodiment of believing that the hand is your own. In other words, this is a direct question of self-identification (Jeannerod & Pacherie, 2004), not just in terms of how participants recognise the movement of the illusory limb (Gallagher, 2000), but also in terms of how they can distinguish between self-generated action and movement on the exterior world (mirror) (Jeannerod, 2006).

The perception that the hand in the mirror is one’s actual unseen hand becomes stronger when there is greater congruence between sensory inputs (embodiment), but the perception becomes weaker when there is a higher incongruity (deafference) (Medina et al., 2015). The mismatch between motor intention/command and actual sensory feedback can also influence embodiment when there is movement during the mirror visual feedback (Jeannerod, 2006). The greater the congruency between the predicted and actual states, the stronger the perception that the illusory hand is one’s actual unseen hand (Moore, 2016). The mismatches between sensory information lead to body representation and ultimately undermine rehabilitation outcomes (Matamala-Gomez et al., 2020).

This study aimed to investigate *believability* over the illusory limb (sense of embodiment) when performing movements with mirror visual feedback. We aimed to examine how this sense was modulated by manipulating four parameters that have been considered important in mirror therapy research (i.e. mirror size, movement execution, task complexity, object manipulation) while participants made movements typical of those used in mirror therapy protocols. Beyond the realism of the mirror image, the sense of embodiment investigates if the hand seen in the mirror is believed to be one’s own hand. By doing so, we sought to address the implications for mirror therapy and illustrate what the optimal mirror therapy conditions might be.

## Methods

### Participants

Forty right-handed (seventeen male; mean age: 21.2 years) participants from the undergraduate student body at the University of Birmingham volunteered to take part in the study. All participants were unimpaired and were naïve to the purpose of the study. The handedness of the participants was self-reported. The study was approved by the University of Birmingham’s Science, Technology, Engineering and Mathematics Ethical Review Committee (ERN_15–1573). Participants provided written informed consent prior to taking part. Recruitment and participation took place between 01/11/2022 and 28/02/2023.

### Apparatus

The experiment was conducted in the Motor Cognition Laboratory, part of the School of Sport, Exercise, and Rehabilitation Sciences at the University of Birmingham. Two different sizes of landscape-oriented Perspex mirrors (50cm X 40cm or 25cm X 20cm) were used depending on the conditions. The mirrors were placed perpendicular to the table and aligned to the participant’s mid-sagittal plane using small bespoke wooden mounts. The large and small mirrors were positioned so that the participant’s dominant hand’s reflection was in view – with the centre of the mirror and their palm in line. The large mirror was fixed with its edge aligned with the table edge, while the small mirror was adjusted to ensure comfort and optimal vision of the reflected limb for each participant. Mirror positions were marked on the table and kept consistent during the experiment. Both hands were placed nine inches away from the mirror to avoid touching the mirror and wooden mounts during the trials (see Figure 1).

**Figure 1.**
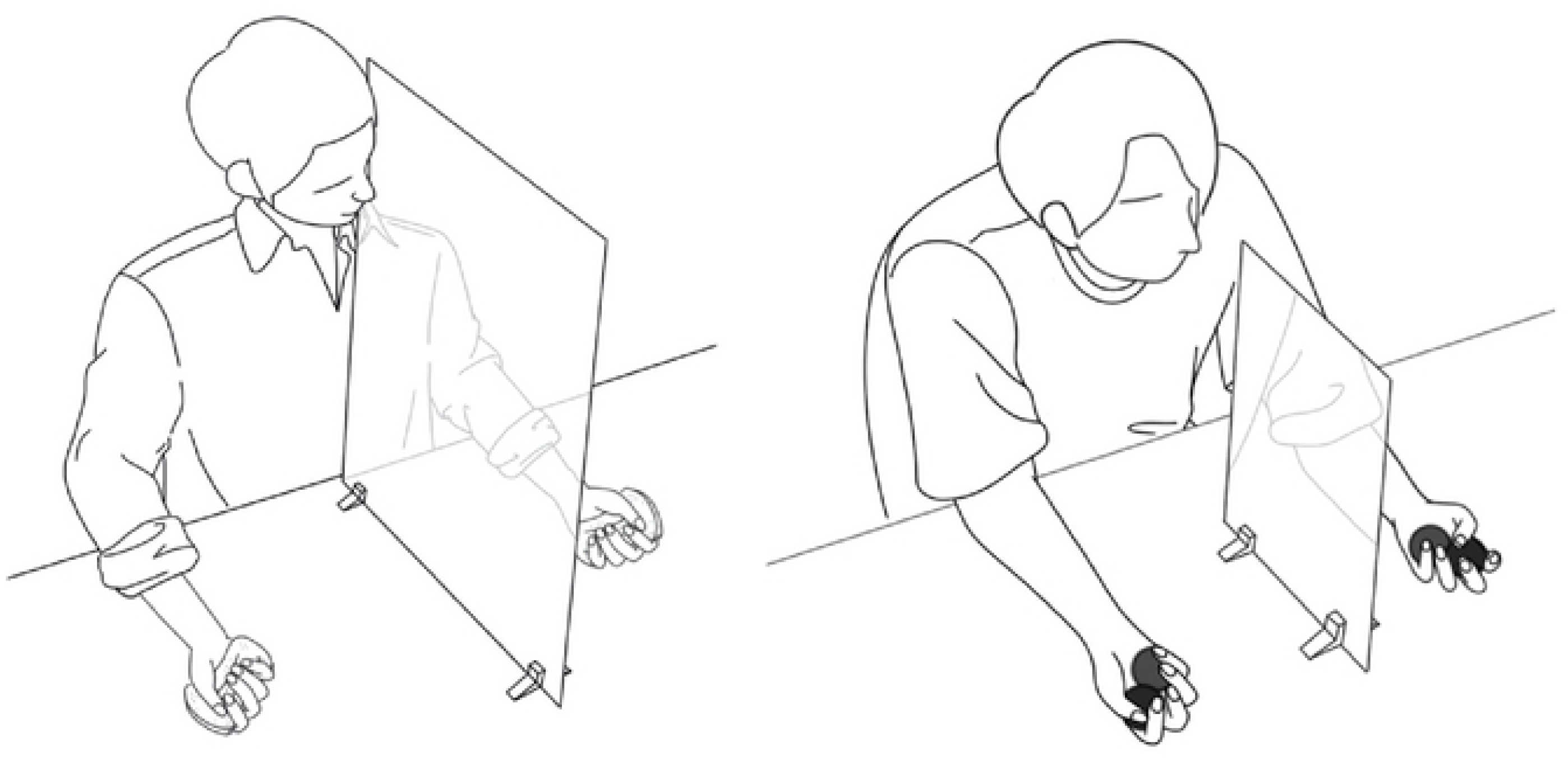
The left figure illustrates one of the conditions in which the participant looks into a large mirror while performing the simple task with manipulating the object. The right figure illustrates one of the conditions in which the participant looks into a small mirror while performing the complex task with manipulating the object. During the trial, participants were directed to look at their hand in the mirror, and both hands were placed 9 inches away from the mirror. Participants completed the task with both hands simultaneously during bimanual execution. However, in unimanual execution conditions, only the hand in front of the mirror (dominant hand) was instructed to move, and the hand behind the mirror did not move with the palm facing up.

### Task, design and procedure

During the study, each trial required participants to perform self-paced repetitive movements for 20 seconds. Participants performed 48 trials during the experiment under 16 conditions; participants completed three trials per condition. Sixteen conditions were created by a combination of four parameters (See Table 1). Details of the parameters are as follows:

(i) Mirror size (large vs. small) Large mirror and small mirror were used depending on the condition.
(ii) Movement execution (unimanual vs bimanual)
  · Unimanual execution: The task was completed with only the dominant (right) hand in front of the mirror, while the unseen non-dominant (left) hand remained static. While performing unimanual execution and also manipulating an object, the object was not held in the unseen hand, which remained static with the palm facing up.
  · Bimanual execution: The hands were instructed to move simultaneously, and while manipulating objects, both hands held objects of the same shape and size.
(iii) Complexity of tasks (simple vs complex) and (iv) manipulation of objects (with vs without object)
  · Simple task without object (see Figure 2-a).
  · Complex task without object (see Figure 2-b).
  · Simple task with object: A sponge (9cm X 4cm X 2cm) was given (see Figure 2-c).
  · Complex task with object: Two wooden balls (2.5cm diameter) were given to each hand (see Figure 2-d).

**Figure 2.**
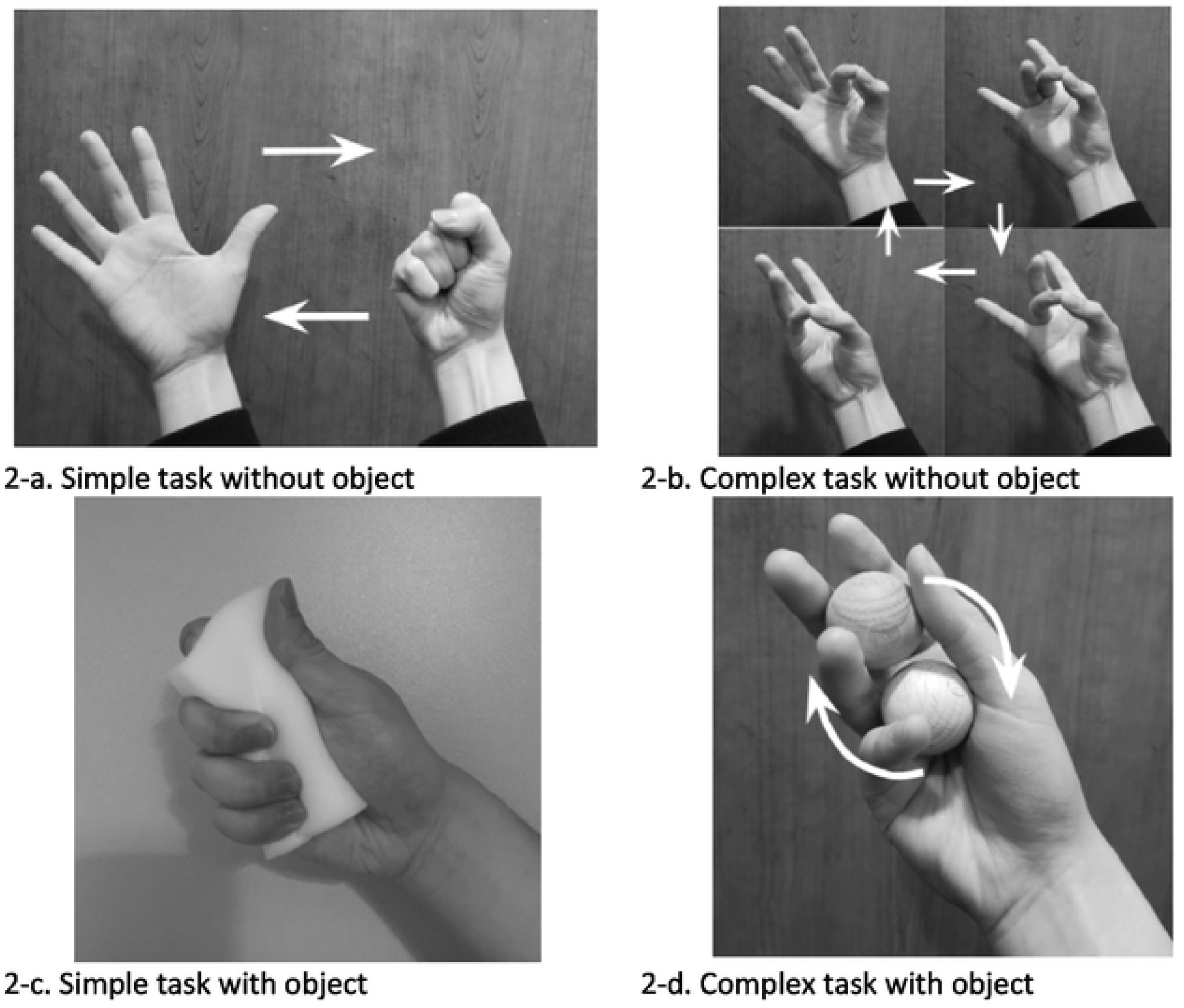
The four conditions combining task complexity and object manipulation parameters. For a ‘simple task without object’, participants were asked to open and close their fist repeatedly (Figure 2-a). For ‘complex task without object’, participants were asked to tap their thumb onto the index, middle, ring and little finger in order and repeat (Figure 2-b). For ‘simple task with object’, participants were asked to squeeze the sponge repeatedly with their palms (Figure 2-c). For ‘complex task with object’, participants were asked to rotate two wooden balls either clockwise with the dominant hand and anti­clockwise with the non-dominant hand (Figure 2-d).

**Table 1.** The sixteen conditions. The conditions were created by a combination of four parameters: (i) mirror size (large vs small), (ii)movement execution (unimanual vs bimanual), (iii)complexity of tasks(simple vs complex), and (iv)object manipulation (with abject vs without object).

|  |  |  |  | Movement execution |  |  |  |
| --- | --- | --- | --- | --- | --- | --- | --- |
|  |  |  |  | Unimanual execution |  | Bimanual execution |  |
|  |  |  |  | Task complexity |  |  |  |
|  |  |  |  | Simple task | Complex task | Simple task | Complex task |
| Mirror size | Large mirror | Object manipulation | Without object | Unimanual simple task without object in large mirror | Unimanual complex task without object in large mirror | Bimanual simple task without object in large mirror | Bimanual complex task without object in large mirror |
|  |  |  | With object | Unimanual simple task with object in large mirror | Unimanual complex task with object in large mirror | Bimanual simple task with object in large mirror | Bimanual complex task with object in large mirror |
|  | Small mirror |  | Without object | Unimanual simple task without object in small mirror | Unimanual complex task without object in small mirror | Bimanual simple task without object in small mirror | Bimanual complex task without object in small mirror |
|  |  |  | With object | Unimanual simple task with object in small mirror | Unimanual complex task with object in small mirror | Bimanual simple task with object in small mirror | Bimanual complex task with object in small mirror |

The presentation of the 16 conditions was randomised across the 48 trials. In all conditions, participants were instructed to direct their vision to the reflection of the hand in the mirror. Fifteen seconds after the start of each trial, the experimenter asked the participants to rate their *believability* on a Likert scale with the following question. “How much do you believe the hand in the mirror feels like your left hand?” The question was answered with a number ranging from zero to ten. Zero representing ‘not at all’, whereas ten represented ‘completely the same’. Once every eight trials, the entire question was posed; the remaining trials only asked for a “please rate from zero to ten” response.

Before commencing the experiment, participants completed the Edinburgh Handedness Inventory and read the written instructions about the procedure of the experiment. Any accessories on the hands and wrists were removed, and any questions regarding the procedure were answered. Participants completed a few practice trials before the experimental trials began in order to familiarise themselves with the procedure. Once experimental trials began, the researcher provided verbal “go” and “stop” signals to indicate the start and finish of trials. Between the trials, there was a short break and a scheduled break after 20 trials.

### Statistical analysis

Data were analysed by using SPSS (IBM SPSS Statistics, Version 28.0. Armonk, NY, USA). The dependent variable of interest was the participant’s *ratings of believability*. The mean ratings for the three repetitions and for each participant were entered for statistical analysis.

Sphericity was verified using Mauchly’s test and statistical values reported accordingly. *Ratings of believability* data were analysed via a 2 × 2 × 2 × 2 (*Mirror* size [large, small] × *Movement execution* [unimanual, bimanual] × *Task complexity* [simple, complex] × *Object manipulation* [with an object, without object]) analysis of variance (ANOVA) with repeated measures. The threshold for statistical significance was set to p < 0.05. Where interactions were found, the simple effects were explored, and Bonferroni correction was applied.

## Results

### Mirror size

The believability was greater when the large mirror (mean = 6.29 ± 2.40) was in place than when replaced by the small mirror (mean = 5.67 ± 2.39), leading to a significant main effect of *Mirror size*, F(1,39) = 34.23, p < 0.001) (see Figure 3-a).

**Figure 3.**
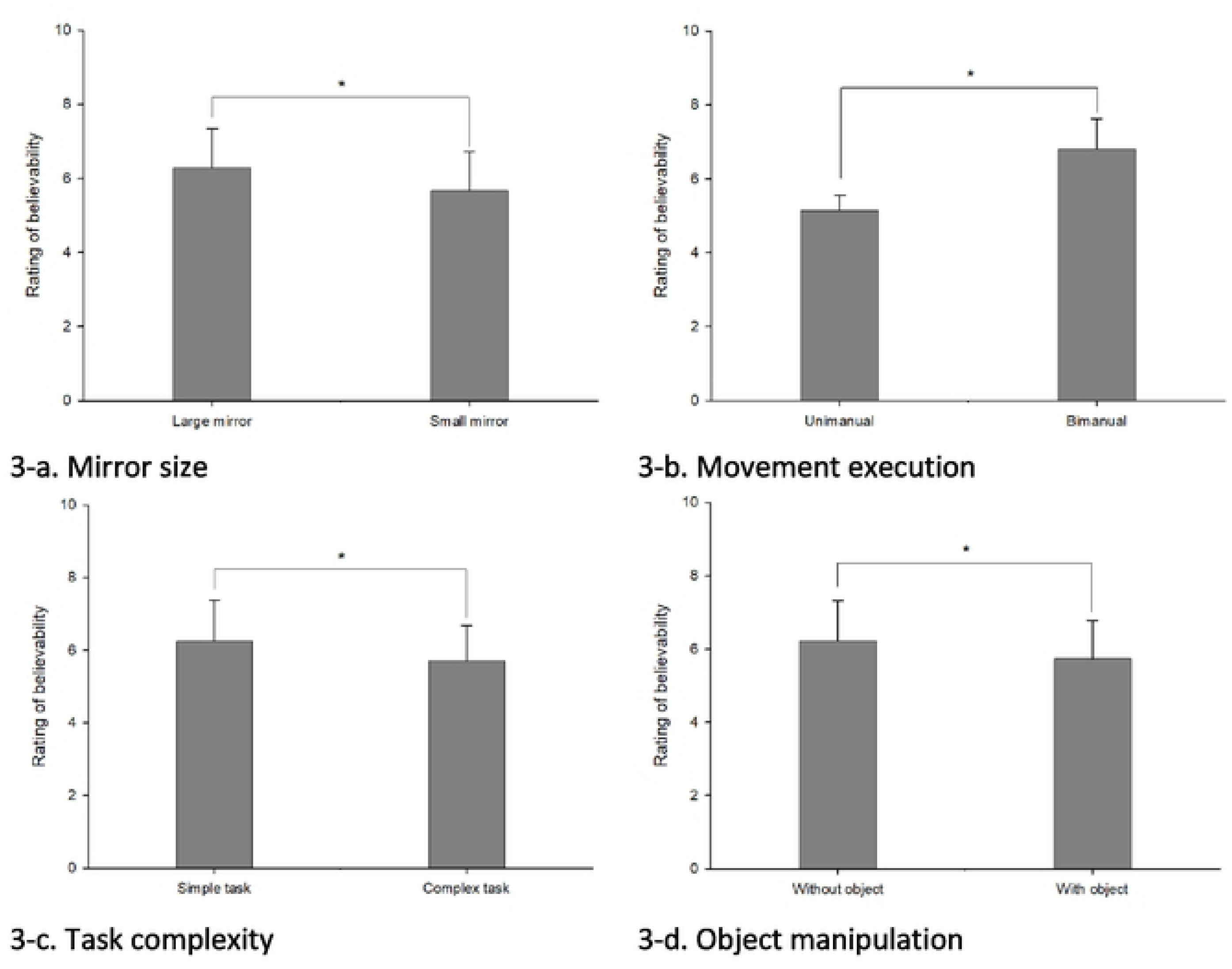
Ratings of believability in each parameter. In this figure, the effects of each parameter are shown by comparing the two levels. The believability was rated out of a total of 10 points. The ratings of believability were shown to be greater for large mirror, bimanual execution, simple task, and when the object was not manipulated.

### Movement execution

The believability was greater when tasks were performed bimanually (mean = 6.81 ± 2.17) rather than unimanually (mean = 5.15 ± 2.36), leading to a significant main effect of *Movement execution*, F(1,39) = 37.85, p < 0.001) (see Figure 3-b).

### Task complexity

The believability was greater when the task was simple (mean = 6.25 ± 2.46) than when the task was complex (mean = 5.71 ± 2.34) leading to a significant main effect of *Task complexity*, F(1,39) = 18.02, p < 0.001) (see Figure 3-c).

### Object manipulation

The believability was greater when the tasks were performed without objects (mean = 6.22 ± 2.38) than when performed with objects (mean = 5.74 ± 2.42) leading to a significant main effect of *Object manipulation*, F(1,39) = 13.31, p < 0.001) (see Figure 3-d).

However, *Movement execution* x *Task complexity*, F(1,39) = 29.07, p < 0.001, *Movement execution* x *Object manipulation*, F(1,39) = 6.78, p = 0.013, *Task complexity* x *Object manipulation*, F(1,39) = 36.15, p < 0.001, and *Movement execution* x *Task complexity* x *Object manipulation*, F(1,39) = 11.17, p = 0.002, interactions suggested a more complex relationship between factors. The results of the three-way interaction are shown in Figure 5.

**Figure 4.**
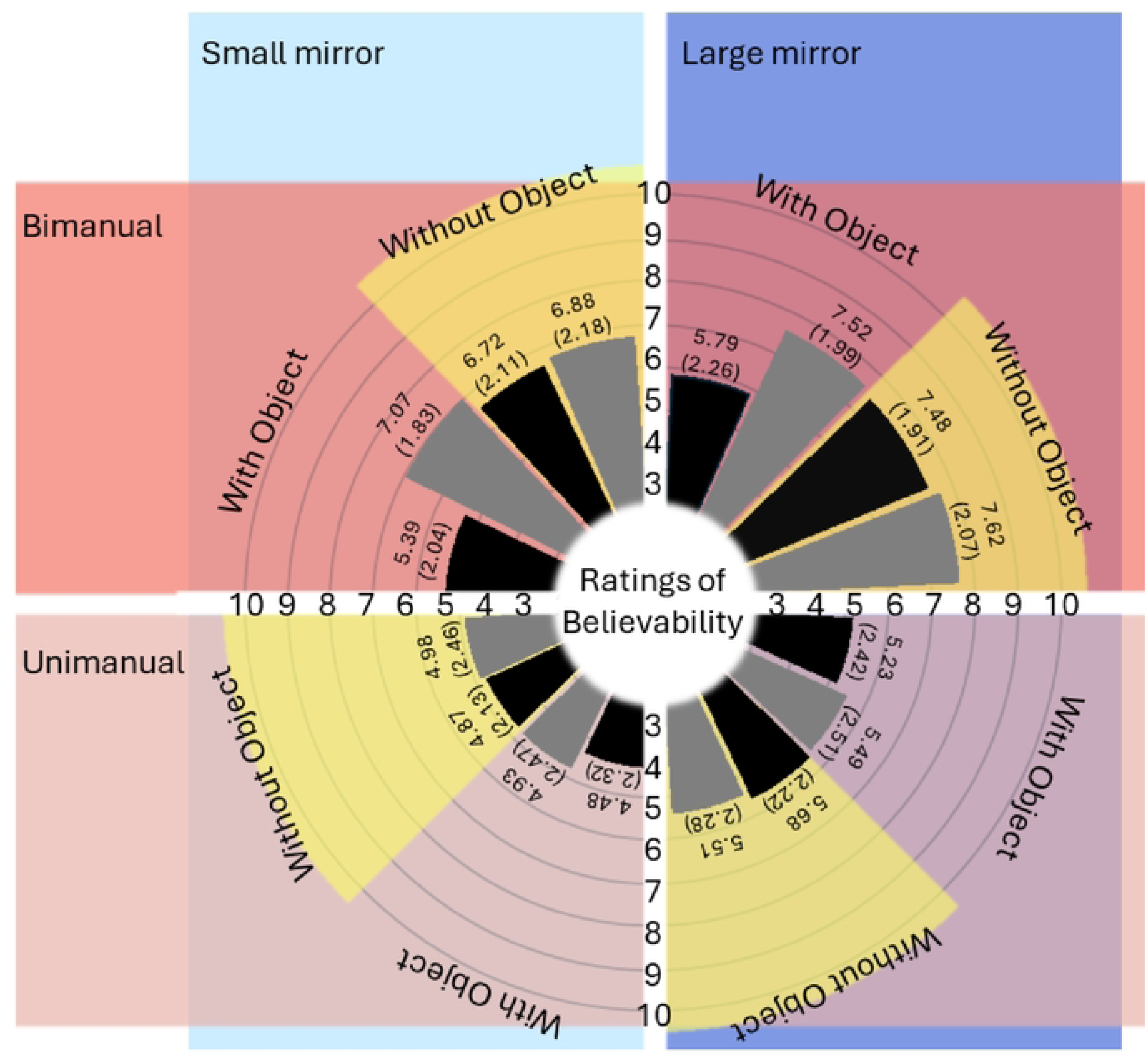
Mean (and standard error) values of believability. The dark blue area represents data collected under the large mirror condition, while the light blue area corresponds to the small mirror condition. The dark red area shows data from bimanual execution, and the light red area displays data from unimanual execution. Four yellow arcs highlight the data collected in the without object condition, distinguishing it from the data related to object manipulation. Black bars represent data from the complex task condition, whereas grey bars depict data from the simple task condition.

**Figure 5.**
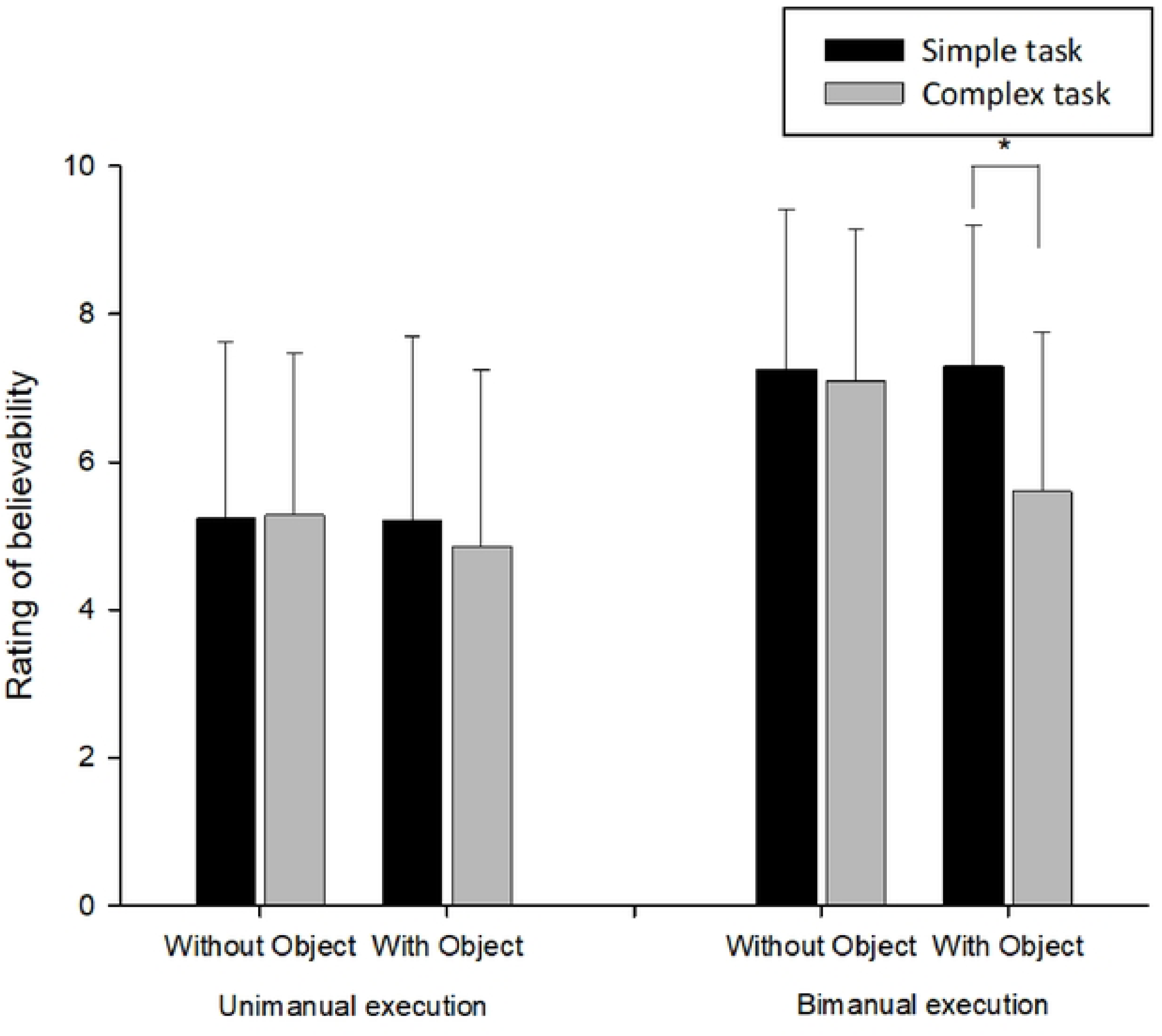
The result of three-way interactions (Movement execution [unimanual, bimanual] x Task complexity [simple, complex] x Object manipulation [with object, without object]). Across the experiment, bimanual execution had higher mean ratings of believability compared to unimanual execution. When tasks were executed unimanually, the mean believability ratings were comparable irrespective of the task complexity and the presence of object used in tasks. However, when tasks were bimanually executed, performing the complex task with object condition significantly decreased the mean ratings of believability.

When performing unimanual movements, the believability between simple and complex tasks with objects, F(1,39) = 3.96, p = 0.054 and without objects, F(1,39) = 0.065, p = 0.800 was comparable. However, when performing bimanual movements, *object manipulation* was responsible for a significant difference between simple and complex tasks, F(1,39) = 31.88, p<0.001. Accordingly, bimanual movements that were complex and required object manipulation resulted in significantly lower average believability (mean = 5.60 ± 2.15) than other bimanual execution conditions (mean = 7.22 ± 2.03). Indeed, the lower ratings in this condition were similar to unimanual condition ratings (mean = 5.15 ± 2.36).

## Discussion

Decades of clinical research have demonstrated the effectiveness of mirror therapy in improving motor function of the hemiparetic limb in stroke survivors (Thieme et al., 2018). However, the intervention is not applied consistently across studies, and the rationale for varying approaches is often unclear (Morkisch et al., 2017). Given the heterogeneity of stroke survivors, a ‘one size fits all’ approach is inadequate, making case-specific modifications essential (McCabe, 2011). Despite this, the optimal protocol for different patient groups remains uncertain.

A recent meta-analysis revealed parameters that resulted in more effective outcomes in hemiparetic stroke (Morkisch et al., 2019). We examined the impact of these parameters on the strength of the resulting illusory experience in unimpaired participants. Consistent with the meta-analysis, it was found that a large mirror elicited a markedly enhanced illusion in comparison to using a small mirror. However, while Morkisch et al. (2019) found unimanual movements were more effective than bimanual movements, we found that bimanual movements were generally responsible for a stronger illusion. Nevertheless, we also found that believability ratings were modified by a combination of factors. More specifically, when unimpaired participants made bimanual movements involving the relatively complex manipulation of objects, the benefits of bimanual movements were lost (i.e. ratings became similar to unimanual movements. Below, these findings are addressed in turn and accounted for, along with what the implications for mirror therapy with stroke might be.

The finding that a large mirror resulted in consistently enhanced believability ratings compared with when a small mirror was in place is consistent with the enhanced effectiveness of mirror therapy demonstrated by the recent meta-analysis (Morkisch et al., 2019). In line with providing what might be considered as a more immersive environment, Rowe et al. (2019) highlights the opportunity afforded by a large mirror to integrate gross muscle movements into a task. This underscores the advantage of a large mirror in reflecting not only the use of the hands, wrists, and forearms but also adequately capturing the movements of the upper arms and shoulders. It allows for the application of tasks that utilise a larger spatial area. McCabe (2011) also highlighted that a large mirror can facilitate a range of bilateral tasks, further expanding its utility. In contrast, a small mirror may limit vision of the illusory limb, and also expose the hidden hand behind the mirror. These factors appear to modulate the quality of the visual illusion created by the mirror and also the effectiveness of mirror therapy.

Several commercially available mirror boxes are small (comparable with the size of the small mirror in this study) offering a limited immersive experience. Instead, the hand is hidden inside the box so that the user may concentrate on their hand in the mirror. However, the enclosed nature of the box may also risk further sensory conflict due to the increased chance of sensory input to the unseen limb (e.g. the hand touching the material that makes up the box sides). Clinical experience suggests patients frequently bump up against the mirror box when moving in limited space available. When this happens, the patient typically pauses the intervention and relocates the position of the unseen hand. This implies that the illusion is disrupted by tactile information gained from touching the box.

In this study, the bimanual execution of movements resulted in generally enhanced believability ratings than those for unimanual movements. As is typical in mirror therapy studies where bimanual movements are employed (Altschuler et al., 1999), participants here made synchronous and symmetrical movements. For unimpaired participants, this clearly results in an experience where one receives visual feedback from the mirror that is congruent with the movements being made with the hidden hand. Under these circumstances, where there is consistency between action and perception, it is perhaps not surprising that bimanual movements result in a more believable experience for participants. In contrast, unimanual movements uncouple action and perception. The findings of Fink et al. (1999) strongly support our explanation. Healthy participants in their study experienced an illusion during out-of-phase movements between the hand in front of the mirror and the hidden hand, highlighting the conflict between visual input from the mirror and proprioceptive feedback from the unseen hand. They reported a heightened sense of peculiarity when sensory information was in conflict rather than congruent, further reinforcing our results.

While predicted, the finding of a more believable illusion when unimpaired participants made bimanual movements (generally at least) is in contrast with Morkisch et al.’s (2017) finding that unimanual movements (i.e. only moving the unimpaired limb) is a more effective approach than bimanual movements when mirror therapy is applied to individuals with stroke. Of course, the measures here are not the same (illusion believability in this study vs. motor improvement for the meta-analysis by Morkisch et al.), but the contrast remains evident. One might speculate that the relative congruence of the behavioural experience in both cases might explain these distinct findings. Where individuals have unimpaired movement, it seems clear that bimanual movements optimise the illusory experience. However, it was also found that making relatively complex movements and manipulating objects reduced this experience to the level of making unimanual movements. Therefore, perhaps any factor that contributes to a lack of congruence between perception and action (Moore, 2016), even where this might be relatively minor, threatens the illusory experience (Fink et al., 1999). For patients with hemiparesis, it might be hypothesised that making bimanual movements provides no greater sense of congruence than making unilateral movements (Swinnen & Wenderoth, 2004). Further, perhaps making bimanual movements also distracts patients from the therapeutic effects of observing the movement in the mirror. While the results of Morkisch et al.’s meta-analysis are unambiguous, it remains possible that the experience may vary for different patients and understanding these relationships more fully would justify further research.

As noted above, believability ratings in the present study for bimanual movements were modulated by task complexity and object manipulation. Previously, Rowe et al. (2019) reported that as the complexity of a task (such as rotating cork balls) increases, the smooth performance of movements may be disrupted. In the present study, the task of rotating the cork balls was defined as difficult. Observations indicated that participants found the task to be challenging, even when performed unimanually. When instructed to rotate the cork balls bimanually, participants struggled to coordinate both hands at the same rate and occasionally dropped the cork balls. Given the task’s difficulty and object to manipulate, successful completion likely required complete focus. The notion that a high perceptual load, which fully utilises the processing capacity for the task, would leave no room for perceiving other input can be applied here (Lavie, 2005). Furthermore, performing the task while looking into a mirror may have exacerbated these challenges, resulting in a discrepancy between the actual perception (actual state) and the anticipated behaviour (predicted state). This, in turn, may have diminished the sense that one has control over the movement of the illusory hand (sense of agency) (Moore, 2016; Synofzik et al., 2008), leading to lower believability ratings.

Interestingly, ratings for bimanual execution did not reduce when combined with just *one* of the other parameters with lower believability (i.e. complexity of task and object manipulation). However, a substantial decline in believability was seen when both parameters were combined, and it appears likely that when ‘overall complexity’ reaches a given threshold it becomes less possible to maintain symmetrical movement of the two limbs and this then threatens the illusion.

As alluded to above, in the case of patients with hemiparesis, even simple movements may be considered ‘complex’ increasing conflict between perception and action; and, depending on the patient, it may simply be preferable not to move the hemiparetic limb (i.e. move the unimpaired limb only). It may also be possible to achieve the desired effect of mirror therapy more optimally by adapting technology. Virtual reality offers an innovative way to overcome the spatial limitations of mirror therapy, enabling patients to experience new environments beyond the confines of traditional approaches (Laver et al., 2017). By providing an immersive and adaptable setting, this technology enhances rehabilitation methods and expands the potential for therapeutic applications (Weber et al., 2019). VR interventions offer enhanced opportunities to incorporate goal-directed tasks and repetitive movements, which are increasingly recognized as essential elements for neurological recovery (Langhorne et al., 2011; Veerbeek et al., 2014).

Another promising possibility for adaptation in mirror therapy is the recently introduced robotic gloves, which could offer a feasible solution. Here, the patient could wear two gloves with the movements of the unimpaired limb being robotically mirrored (and controlled) by a glove worn on the impaired limb. In this scenario, it should be possible to create conditions where patients see and feel movements with their impaired limb that perfectly match those of the unimpaired limb. And while the movement of the impaired limb may be passive, proprioceptive signals will still be produced, likely enhancing the perceptual experience (Proske & Gandevia, 2012).

## Conclusion

In summary, we examined the impact of different parameters on the subjective strength of the illusion elicited by mirror therapy in unimpaired individuals, by measuring to what extent participants *believed* that the hand in the mirror was their *unseen* hand. Large mirrors elicited higher ratings than small mirrors, and bimanual execution elicited higher ratings than unimanual execution. However, when bimanual execution was combined with a complex task and object manipulation, the believability ratings were markedly lower (comparable with unimanual execution). Overall, findings are consistent with the importance of maintaining congruency between perception and action in order to optimise the illusory experience that is the aim of mirror therapy. Task difficulty threatens this congruence and careful consideration should be paid to the details of the mirror therapy procedure depending in the abilities of individual patients.

## Data Availability

Due to the sensitive nature of the data, information created during and/or analysed during the current study is available from the corresponding author (Jin Min Kim/) on reasonable request to bona fide researchers.

